# Regression Models for Predictions of COVID-19 New Cases and New Deaths Based on May/June Data in Ethiopia

**DOI:** 10.1101/2020.09.04.20188094

**Authors:** Alemayehu Siffir Argawu

## Abstract

As the 15 of June 2020, we have 7,984,067 total COVID-19 cases, globally and 435,181 total deaths. Ethiopia was ranked 2^nd^ and 15^th^ in the table by 176 new cases and by 3,521 total new cases from African countries. Then, this study aimed to predict COVID-19 new cases and new deaths based on May/June data in Ethiopia using regression model. In this study, I used Pearson’s correlation analysis and the linear regression model to predict COVID-19 new cases and new deaths based on the available data from 12^th^ May to 10^th^ June 2020 in Ethiopia. There was a significant positive correlation between COVID-19 new cases and new deaths with different related variables. In the regression models, the simple linear regression model was a better fit the data of COVID-19 new cases and new deaths than as compared with quadratic and cubic regression models. In the multiple linear regression model, variables such as the number of days, the number of new laboratory tests, and the number of new cases from AA city significantly predicted the COVID-19 new cases. In this model, the number of days and new recoveries significantly predicted new deaths of COVID-19. The number of days, daily laboratory tests, and new cases from Addis Ababa city significantly predicted new COVID-19 cases, and the number of days and new recoveries significantly predicted new deaths from COVID-19. According to this analysis, if strong preventions and action are not taken in the country, the predicted values of COVID-19 new cases and new deaths will be 590 and 12 after two months (after 9^th^ of August) from now, respectively. The researcher recommended that the Ethiopia government, Ministry of Health and Addis Ababa city administrative should give more awareness and protections for societies, and they should also open more COVID-19 laboratory testing centers. Generally, the obtained results of this study may help Ethiopian decision-makers put short-term future plans to face this epidemic.

## 1. Introduction

Corona virus disease (COVID-19) is an infectious disease that is caused by severe acute respiratory syndrome known as corona virus. The virus was first identified on 31 December 2019 in the city of Wuhan, which is the capital of Hubei Province in China. Some of the common signs of COVID-19 include fever, shortness of breath and dry coughs. Other uncommon symptoms include muscle pain, mild diarrhea, abdominal pain, sputum production, loss of smell, and sore throat. On 11 of March 2020, the WHO announced that it was a global pandemic [1–5].

On 15^th^ of June 2020, as the Worldometer corona virus updates information reported that we have 8,028,253 total COVID-19 cases, 436,276 total deaths with 5.4% fatality rate and 4,148,128 total recovered with 51.7% recovered rate as globally. It was distributed from highest to lowest ranks of the new cases and deaths by the World Regions as follows: North America has led by 2,480,701 total new cases (1^st^) and 144,979 total deaths (2^nd^), Europe has 2,398,779 total new cases (2^nd^) and 188,001 total deaths (1^st^), Asia has 1,616,962 total new cases (3^rd^) and 40,248 total deaths (4^th^), South America has 1,425,696 total new cases (4^th^) and 60,457 total deaths (3^rd^), Africa has 244,578 total new cases (5^th^) and 6,490 total deaths (5^th^), and the last in both ranks is Oceania, which has 8,931 cases and 124 deaths. In the report, the male and female cases were 71% and 29%, respectively [6].

On this date, Ethiopia was ranked as the 2^nd^, 15^th^, 16^th^ and 23^rd^ on the table by 176 new cases, by 3,521 total COVID-19 cases, by 60 total deaths (1.7%) and by 620 total recovered (17.6%) as compared from African countries as the Worldometer corona virus updates information showed [6].

The report also showed that Ethiopia was listed on the 27^th^ place by the capacity of COVID-19 laboratory tests. It was 1,629 tests per 1,000,000 populations. This is bad news for Ethiopia. Currently, the Ethiopian population is near 115 million. This is the fact that Ethiopia has a very low proportion of COVID-19 laboratory tests compared with other countries tests. This report indicated that Ethiopia needs increasing efforts and strategies to increase daily laboratory tests. Otherwise, she will be the next ‘‘African USA’’. Then, this study aimed to achieve the following objectives:

1. To predict COVID-19 new cases based on the number of days, and daily laboratory tests using a regression model.
2. To predict new deaths based on the number of days, new cases, and new recovered using a regression model.

## 2. Method

### 2.1. Data

The COVID-19 new case report data were collected from Ethiopia Ministry of Health and Ethiopian Public Heath Institution reports from their face book and telegram pages [7, 8]. 14^th^ of March 2020 was the first date that COVID-19 was confirmed in Ethiopia. The time period of data was from the 12^th^ of May to the 10^th^ of June 2020 (for the last 30 days). The data included the total number of new cases, date of recorded, number of new total COVID-19 cases, number of new deaths, number of new recoveries, persons who have contacted infected cases, number of male total COVID-19 cases, number of new cases from AA city, and others.

In this study, I used Pearson’s correlation analysis and the linear regression model to predict COVID-19 new cases based on the available data from 12^th^ May to 10^th^ June 2020 in Ethiopia.

### 2.2. Regression Model

Regression analysis is a statistical technique for investigating and modeling the relationship between variables. Applications of regression are numerous and occur in almost every field, including engineering, life and biological sciences, and other sciences. In fact, regression analysis may be the most widely used statistical technique.

The regression model has many variants such as linear regression, polynomial regression, and others [9].

### 2.2.1. Linear Regression Model

The fitted multiple linear regression model was used to determine the most predicator variables for COVID-19 new cases and new deaths from 12^th^ May to 10^th^ June 2020 in Ethiopia.

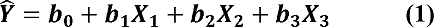

Where: Y is COVID-19 new cases or new deaths. And X_1_, X_2_, and X_3_ are independent variables with their corresponding coefficients (b_1_, b_2_, and b_3_). And, b_0_ is the intercept coefficient in the model.

### 2.2.2. Polynomial Regression Model

The fitted cubic regression model was used to estimate the parameters of independent variables as X, X^2^, and X^3^. All the estimated parameters (b_1_, b_2_, and b_3_) shown the change of Y when the independent variable changed from x to x+1.

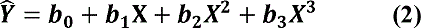

Where: Y is COVID-19 new cases or new deaths. And X is an independent variable with corresponding regression coefficient b(s). And, b_0_ is the intercept coefficient in the model.

## 3. Result

### 3.1. Frequency Statistics of COVID-19 Cases

#### Total number of COVID-19 laboratory tests

The total numbers of COVID-19 laboratory tests were 158,521 and 121,938 from 14^th^ of March to 10^th^ of June and 12^th^ of May to 10^th^ of June, respectively. This indicates that the testing rate was increased to 77%. The newly reported COVID-19 laboratory tests peaked at 6,092 on June 07, 2020.

#### Prevalence of COVID-19 cases

The total COVID-19 cases were 2,506 and 2,257 from 14^th^ of March to 10^th^ of June and from 12^th^ of May to 10 of June, respectively. This indicates that new cases from 12^th^ May to 10^th^ June (duration of 30 days) were *increased by 10 times* from 14^th^ March to 11^th^ May (duration of 60 days). Thus, the prevalence of COVID-19 new cases increased from 158 to 185 per 10,000 laboratory tests per day from 12^th^ May to 10^th^ June in Ethiopia, 2020. The new cases peaked at 190 cases on June 09, 2020.

#### Death rate

The crude mortality or death rate was 1.4% (35) and 1.3% (30) from 14 to 10 June and from 12 May to 10 June, respectively. This showed that the death of COVID-19 was *6 times greater from* 12^th^ May to 10^th^ June than from 14 March to 11 May in Ethiopia, 2020. New deaths peaked at 7 deaths on June 7^th^ in 2020.

#### Recovery rate

The recovery rates were 16% (401) and 13% (294) from 14 March to 10 June and from 12 May to 10 June, respectively. In addition, 73.3% of the total recovered cases were reported from 12th May to 10th June in Ethiopia, 2020.

**Table 1.** Percentage and rates of COVID-19 cases from 14^th^ of March to 10 of June and from 12^th^ May to 10^th^ June in Ethiopia, 2020

| No. | Variable | Date |  |
| --- | --- | --- | --- |
|  |  | 14<sup>th</sup> March to 10<sup>th</sup> June (90 days) | 12<sup>th</sup> May to 10<sup>th</sup> June (30 days) |
| 1. | Daily laboratory tests | 158, 521 | 121, 938 (77%) |
| 2. | COVID-19 new cases | 2, 506 (1.58%) | 2, 257 (1.85%) |
| 3. | New deaths | 35 (1.4%) | 30 (1.3%) |
| 4. | New recoveries | 401 (16%) | 294 (13%) |

### 3.2. COVID-19 New Cases by Regions and Genders

Table 2 below illustrates the percentages of COVID-19 cases by gender and region from 12th May to 10th June (for the last 30 days) in Ethiopia, 2020.

From the total number of 2,257 COVID-19 new cases, the majorities (64%) were males and 36% of them were females. This indicated that male group was infected less in Ethiopia as compared with world male cases was 71% [6]. Addis Ababa city has covered the majority (74%) of the Pandemic. The Somali region (7%) has taken the 2^nd^ highest coverage of the virus. Oromia and Amhara regions have equally shared the COVID-19 new cases (each 6%). However, Tigray and other regions (such as SNNP, Afar, Harare, etc.) have taken only 3% of COVID-19 new cases by each. In addition, foreign natives have received only 1% of the distribution.

From the history of infected cases, patients’ contacts and travel histories not known accounted for the majority (54%, 594) of the COVOD-19 total COVID-19 cases. The patients’ travel history from abroad and contacts with other new cases covered 26% (283) and 20% (218), respectively. All these histories of the cases were considered from May 12^th^ to June 2^nd^, 2020.

**Table 2.** Prevalence of COVID-19 new cases by gender and region from 12^th^ May to 10^th^ June (for the last 30 days) in Ethiopia, 2020

| No. | Prevalence by Category | # of days | Total | % |
| --- | --- | --- | --- | --- |
| 1. | Gender |  | 2,257 | 100 |
|  | Male | 30 | 1,441 | 64 |
|  | Female | 30 | 816 | 36 |
| 2. | Region |  | 2,237 | 99 |
|  | Addis Ababa City | 30 | 1,671 | 74 |
|  | Oromia Region | 30 | 130 | 6 |
|  | Amhara Region | 30 | 134 | 6 |
|  | Tigray Region | 30 | 78 | 3 |
|  | Somali Region | 30 | 157 | 7 |
|  | Other Regions | 30 | 67 | 3 |
|  | Other (Foreign Natives) | 30 | 20 | 1 |
| 3. | Patient's History |  | 1,095 | 100 |
|  | Case had travel history from abroad | 22 | 283 | 26 |
|  | Case had contact with confirmed case | 22 | 218 | 20 |
|  | Case hadn't known contact & travel history | 22 | 594 | 54 |

### 3.3. Descriptive Statistics of COVID-19 Cases

Table 3 below demonstrates the descriptive statistics of the COVID-19 new cases from 12th May to 10th June in Ethiopia, 2020.

**Table 3.** Descriptive statistics of the COVID-19 cases from 12^th^ May to 10^th^ June (for the last 30 days) in Ethiopia, 2020

| No. | Variables | Min. | Max. | Mean | S.D |
| --- | --- | --- | --- | --- | --- |
| 1. | Daily laboratory tests | 1775 | 6,187 | 4,065 | 1,094 |
| 2. | COVID-19 new cases | 2 | 190 | 75 | 57 |
|  | New cases from Addis Ababa city | 0 | 153 | 56 | 48 |
| 3. | Minimum age of new cases | 0.1 | 24 | 9.4 | 6.9 |
| 4. | Maximum age of new cases | 33 | 115 | 71 | 18 |

#### Daily laboratory tests

The average value of COVID-19 conducted laboratory tests was 4,065 per day with its min (1,775) and max (6,187) in the given duration.

#### COVID-19 new cases

The average value of COVID-19 new cases was 75 per day, with minimum and maximum values of 2 and 190, respectively. AA city had recorded the highest COVID-19 new cases (56) per day in the given duration.

The average values of the minimum and maximum ages of COVID-19 new cases were 9.4 years and 71 years with their smallest and largest ages of 1 month and 115 years, respectively.

### 3.4. Correlation Analysis for COVID-19 New Cases

The correlation analysis showed that there were significantly positive correlations between COVID-19 new cases and the number of days, daily laboratory tests, new cases of males, new cases of females, new cases from AA city, new cases from foreign natives, new cases with unknown contact and travel histories, and new cases with contact with infected persons (table 4).

**Table 4.**
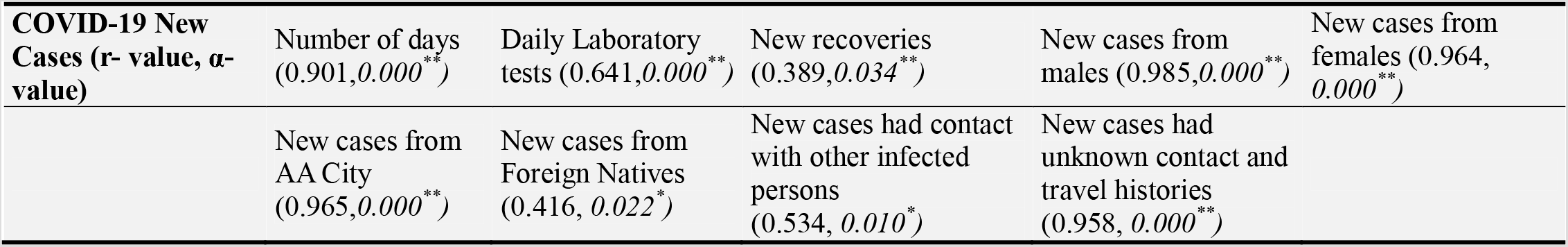
Correlation analysis between COVID-19 New Cases and related variable

### 3.5. Correlation Analysis for New Deaths

Additionally, the correlation analysis for new deaths showed that there were significantly positive correlations between the number of new deaths and the number of days, COVID-19 new cases, daily laboratory tests, new cases from AA city, new recoveries, new cases of males, new cases of females and maximum age of new cases. However, there was a significantly negative correlation between new deaths and the minimum age of new cases (Table 5).

**Table 5.**
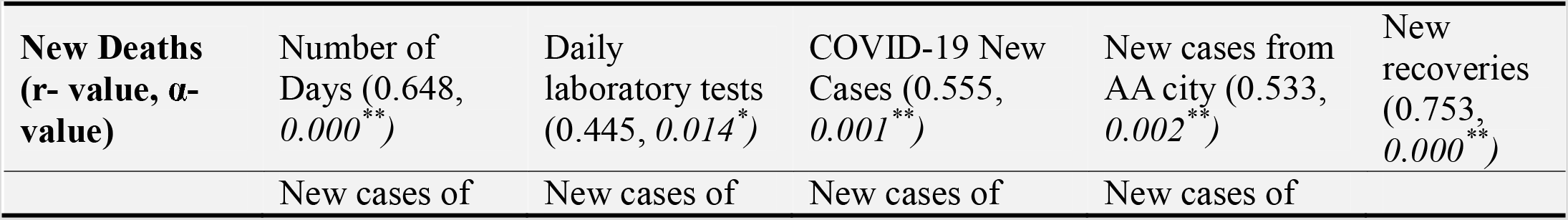
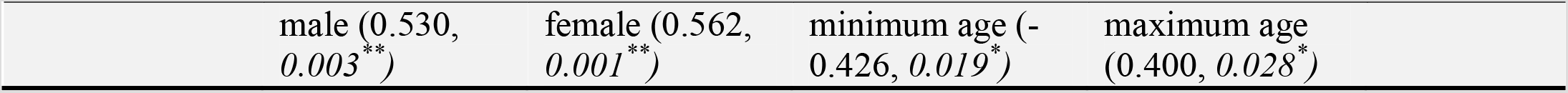
Correlation analysis for the relationship b/n New Deaths of COVID19 and related variables

### 3.6. Regression Model for COVID-19 New Cases

Table 6 showed that the linear regression model has the highest F-value (120.7) and the smallest MSE value (637.4) as compared with quadratic and cubic models. And, the number of days was significant predictor for new cases in the linear regression model (p-value of 0.000). But, this variable and its two expressions were not predictors in the quadratic and cubic regression model.

**Table 6.** Regression model summary and parameter estimates to predict COVID-19 New Cases

| Independent Variable | Model | R<sup>2</sup> | F | MSE | Sig. | Constant | b1 | b2 | b3 |
| --- | --- | --- | --- | --- | --- | --- | --- | --- | --- |
| Day | Linear | 0.81 | **120.7** | **637.4** | 0.000 | -378.2 (**0.000**)<sup>*</sup> | 5.851 (**0.000**)<sup>*</sup> |  |  |
|  | Quadratic | 0.82 | 59.8 | 646.3 | 0.000 | -55.7(0.894) | -2.577(0.813) | 0.054(0.440) |  |
|  | Cubic | 0.82 | 59.8 | 646.3 | 0.000 | -55.7(0.894) | -2.577(0.813) | 0.054(0.440) | 0 |
| Daily Laboratory Tests | Linear | 0.41 | **19.5** | 1993.3 | 0.000 | -61.0(**0.000**)<sup>*</sup> | 0.034(**0.000**)<sup>*</sup> |  |  |
|  | Quadratic | 0.43 | 10.4 | 1983.8 | 0.000 | 37.0(0.707) | -0.017(0.722) | 0 |  |
|  | Cubic | 0.46 | 7.5 | 1952.2 | 0.001 | 338.8(0.220) | -0.273(0.223) | 0.0001(0.204) | 0 |
| Addis Ababa City New Cases | Linear | 0.93 | **382.6** | **230.8** | 0.000 | 10.8 (**0.000**)<sup>*</sup> | 1.2 (**0.000**)<sup>*</sup> |  |  |
|  | Quadratic | 0.93 | 187.4 | 235.9 | 0.000 | 9.1(0.090) | 1.3 (**0.000**)<sup>*</sup> | -0.001(0.533) |  |
|  | Cubic | 0.93 | 122.3 | 241.2 | 0.000 | 7.02 (0.262) | 1.6(**0.006**)<sup>*</sup> | -0.007(0.469) | 0 |

\* The regression coefficients are significant at 1%.

**Table 6.** Regression model summary and parameter estimates to predict New Deaths

| Independent Variable | Model | R<sup>2</sup> | F | MSE | Sig. | Constant | b1 | b2 | b3 |
| --- | --- | --- | --- | --- | --- | --- | --- | --- | --- |
| Day | Linear | 0.42 | **20.2** | **1.7** | 0.000 | -8.6 (**0.000**)* | 0.12 (**0.000**)* |  |  |
|  | Quadratic | 0.48 | 12.7 | 1.6 | 0.000 | 28.8 (0.169) | -0.85 (0.119) | 0.01(0.076) |  |
|  | Cubic | 0.48 | 12.7 | 1.6 | 0.000 | 16.4 (0.243) | -0.37(0.182) | 0 | 0 |
| New Cases | Linear | 0.31 | **12.5** | **2.02** | 0.001 | -0.23 (0.602) | 0.016 (**0.001**)* |  |  |
|  | Quadratic | 0.31 | 6.1 | 2.1 | 0.007 | -0.12 (0.842) | -0.012 (0.499) | 0 |  |
|  | Cubic | 0.36 | 4.6 | 2.1 | 0.011 | -0.76 (0.353) | -0.061 (0.191) | -0.001(0.278) | 0 |
| New Recovered | Linear | 0.57 | **36.7** | **1.27** | 0.000 | -0.02 (0.935) | 0.10 (**0.000**)* |  |  |
|  | Quadratic | 0.58 | 18.4 | 1.29 | 0.000 | 0.14 (0.681) | 0.07 (0.103) | 0.001 (0.453) |  |
|  | Cubic | 0.58 | 11.9 | 1.33 | 0.000 | 0.23 (0.613) | 0.03 (0.817) | 0.003(0.759) | 0 |

\* The regression coefficients are significant at 1%.

Thus, the fitted linear regression model was much better than the quadratic and cubic models.

However, the three models have a similar R square values as 81% and 82% variations of COVID-19 new cases was explained by the models (figure 1). Then, the estimated linear regression equation is

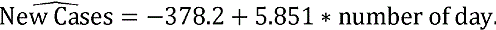

**Figure 1:**
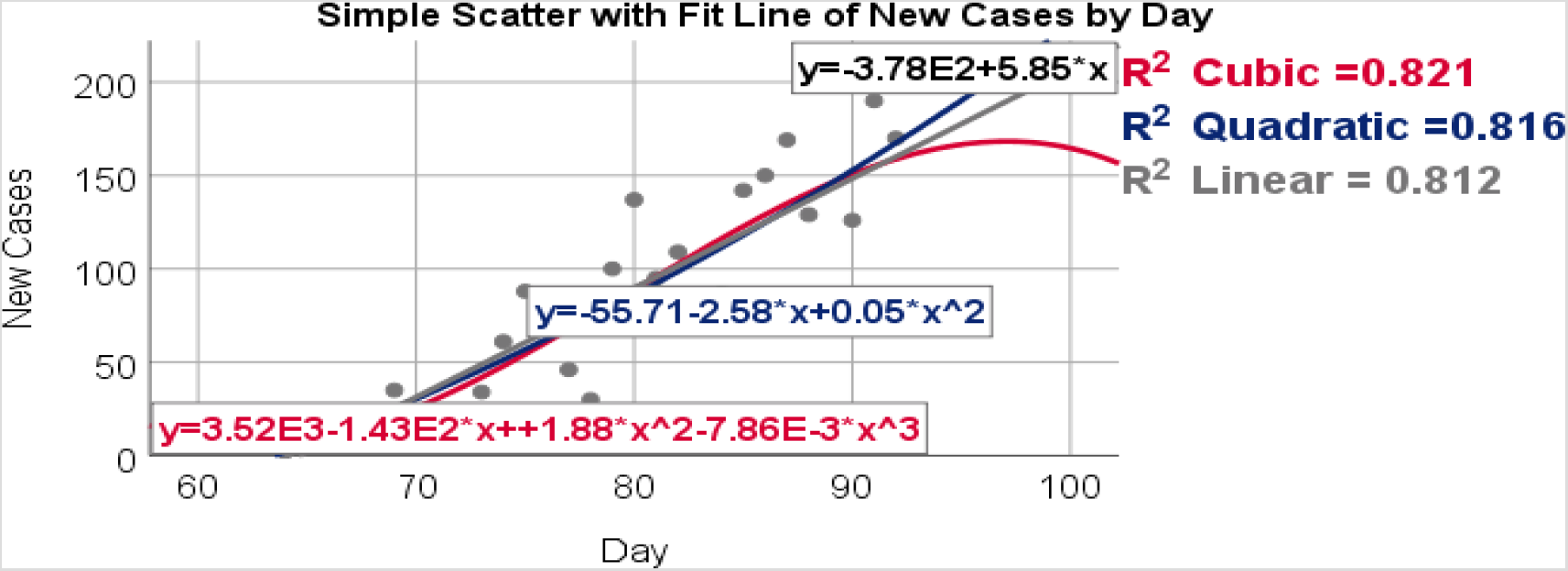
Simple line graph of COVID-19 new cases by the number of days

This implied that the new cases will be increased to 585 after 100 days.

And the table also showed that a daily laboratory test was significant predictor for new cases in the linear regression model (p-value of 0.000). But, this variable and its two expressions were not predictors in the quadratic and cubic regression model.

And, the fitted linear regression model has the highest F-value (19.5) and but not the smallest MSE value (1993.3) as compared with quadratic and cubic models. Thus, the fitted linear regression model was much better than the quadratic and cubic models.

However, the cubic regression model has a better R square value as 46% variations of COVID-19 new cases was explained by the model. And, the linear regression model explained 41% of the variations (figure 2). From the table, the fitted linear regression equation is

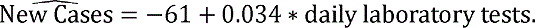

**Figure 2:**
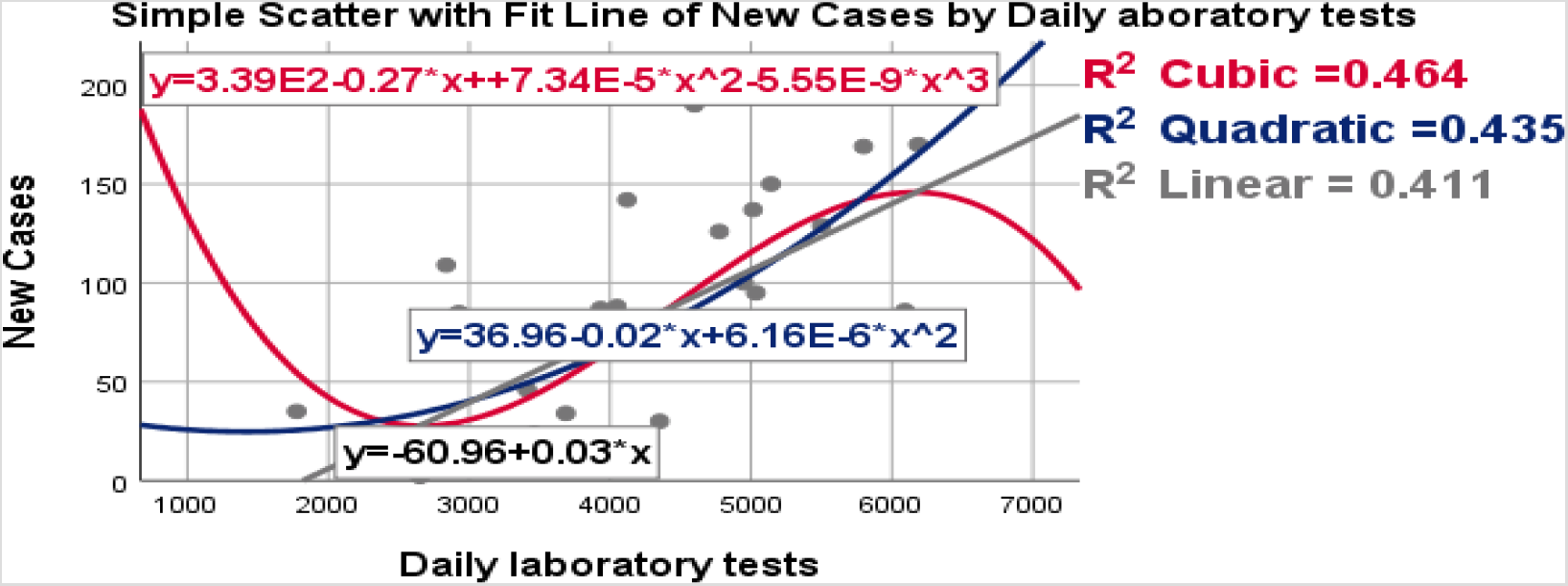
Simple line graph of COVID-19 new cases by daily laboratory tests

This reported that the new cases will be raised to 3,400 if 100,000 laboratory tests were conducted daily. Similarly, new case from Addis Ababa city was predicted significantly the new cases in the linear regression model (p-value of 0.000) with R^2^ = 93% of the new cases variations was explained by cases from Addis Ababa city (figure 3).

Then, the estimated linear regression equation is

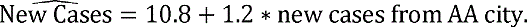

**Figure 3:**
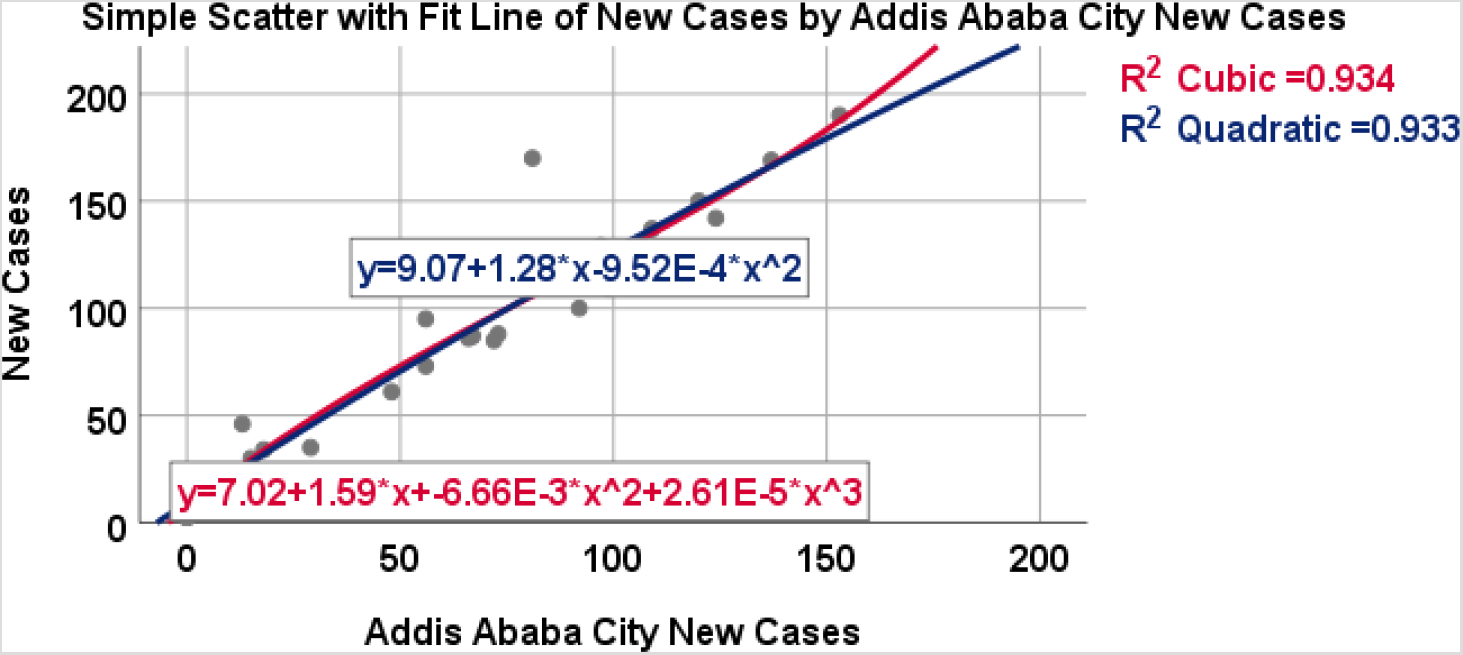
Simple line graph of COVID-19 new cases by new cases from AA city

This suggested that the country’s new cases will be increased to 12,000 if 10,000 new cases were found in AA city.

### 3.7. Regression Model for New Deaths

Table 7 presented that the linear regression model has the highest F-value (20.2) and the smallest MSE value (1.7) as compared with quadratic and cubic models. And, the number of days was significant predictor for new deaths in the linear regression model (p-value of 0.000). But, this variable and its two expressions were not predictors in the quadratic and cubic regression model.

Thus, the fitted linear regression model was much better than the quadratic and cubic models.

However, the quadratic or cubic models have a better R^2^ = 48% and the linear regression model has R^2^ = 42% of variation of new death was explained by the models (figure 4). Then, the estimated linear regression equation is

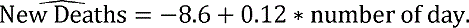

**Figure 4:**
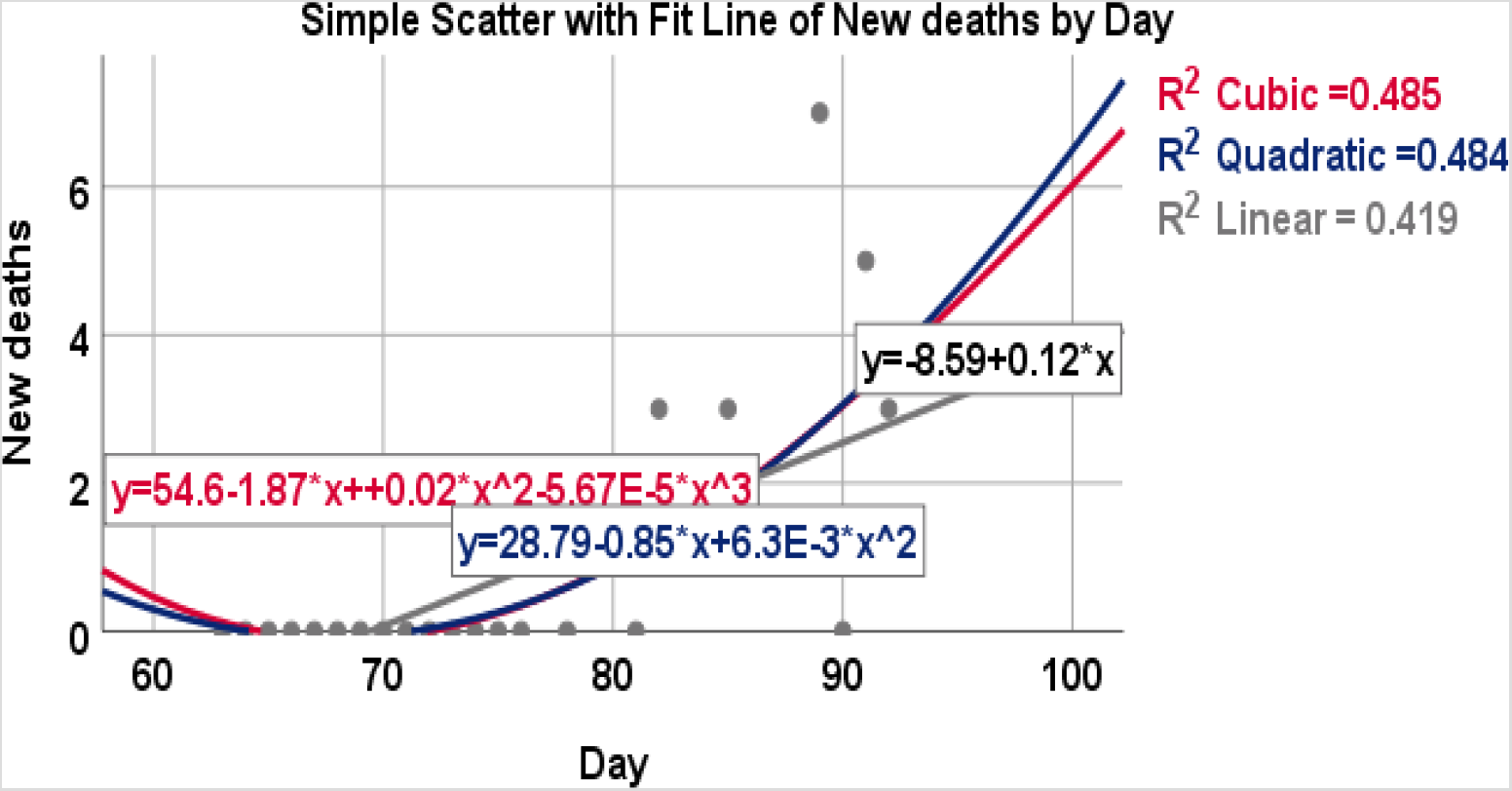
Simple line graph of COVID-19 new deaths by the number of days

This suggested that the new deaths will be increased to 12 after 100 days.

And the table also showed that a new case was significant predictor for new deaths in the linear regression model (p-value of 0.001). And, the fitted linear regression model has the highest F-value (19.5) and the smallest MSE value (2.02) as compared with quadratic and cubic models. Thus, the fitted linear regression model was much better than the quadratic and cubic models.

However, the cubic regression model has a better R square value as 36% variations of COVID-19 new deaths was explained by the model. And, the linear regression model explained 31% of the variations (fig 5). From the table, the fitted linear regression equation is

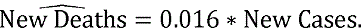

**Figure 5:**
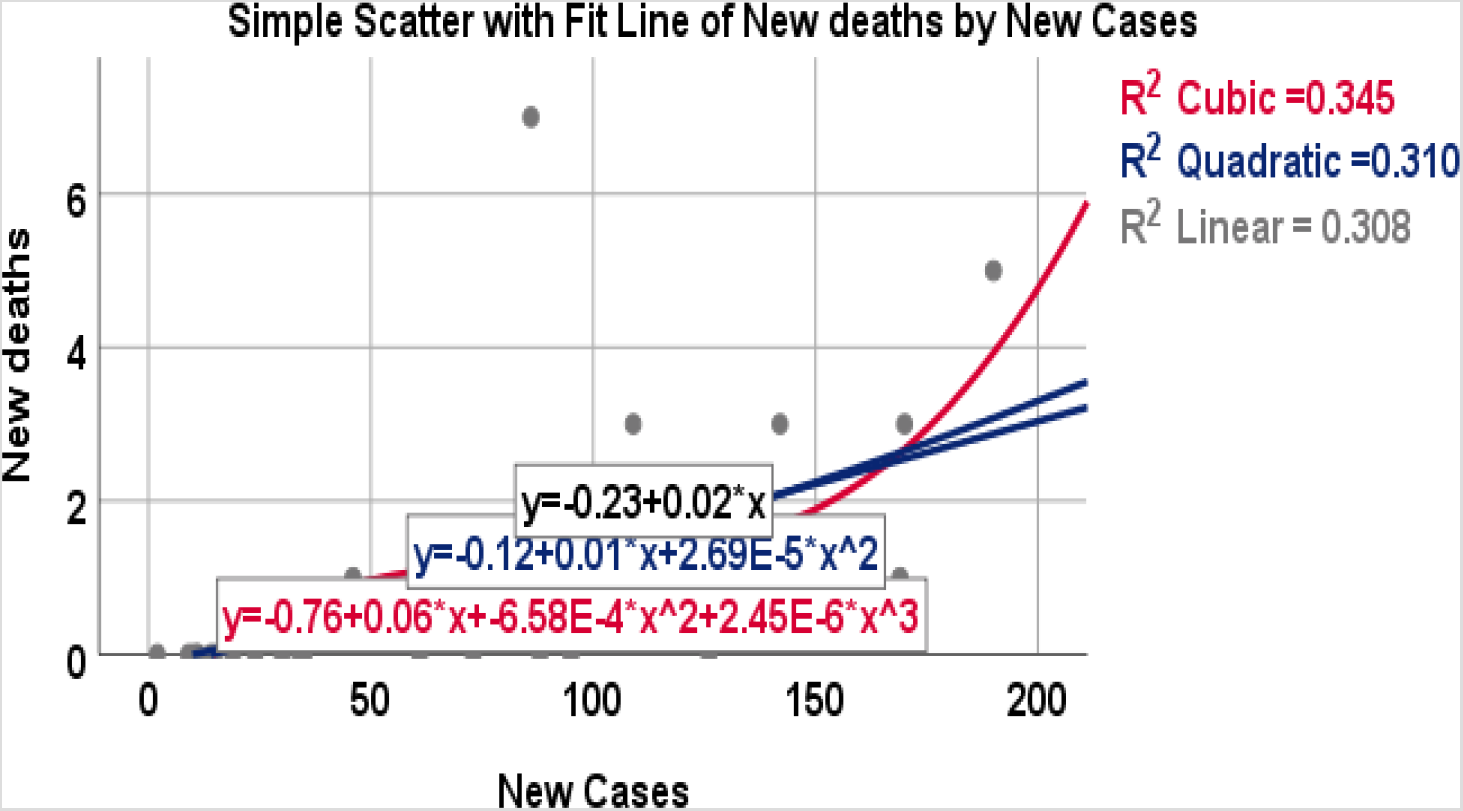
Simple line graph of new deaths by new cases

This indicated that the new deaths will be raised to 16 if 1,000 new cases were found.

Similarly, new recovered was predicted significantly the new deaths in the linear regression model (p-value of 0.000) with R^2^ = 57% of new deaths variations was explained by new recovered (figure 6).

Then, the estimated linear regression equation is

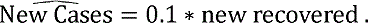

**Figure 6:**
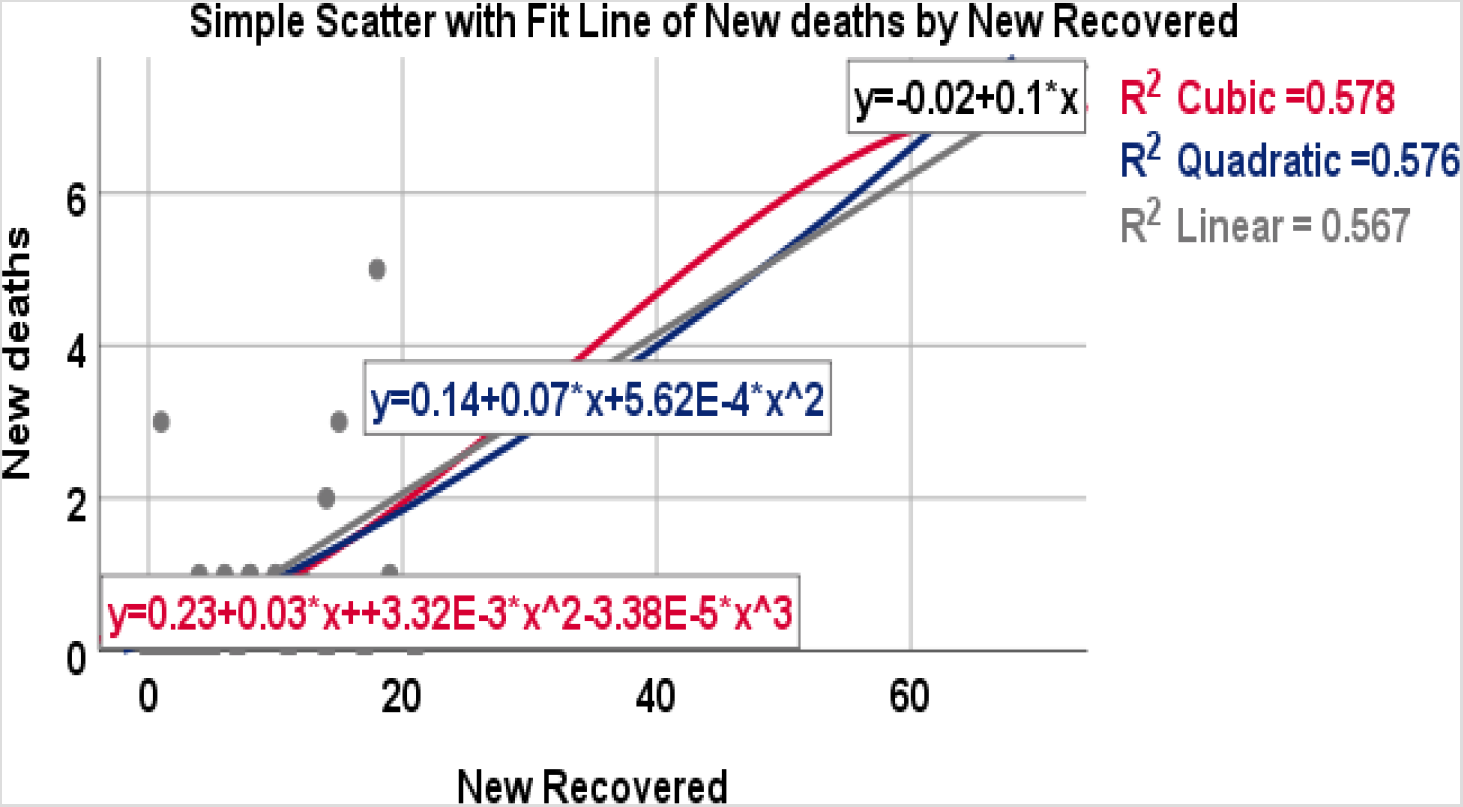
Simple line graph of new deaths by new recovered

This implied that the new deaths will be increased to 100 if 1,000 patients recovered from the virus.

### 3.8. Multiple Linear Regression Model for COVID-19 New Cases

In this model, COVID-19 new cases were predicted significantly by the number of days, daily laboratory tests and new cases from Addis Ababa city at the 5%, 10% and 1% levels of significance, respectively.

The fitted MLR for COVID-19 New Cases,

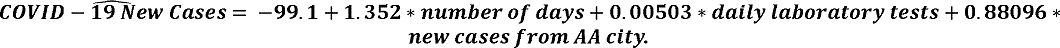

1. COVID-19 new cases are predicted to increase 135 when the number of days increases by 100 days while holding other variables constant.
2. COVID-19 new cases are predicted to increase 503 when the daily laboratory tests rise by 100,000 tests while holding other variables constant.
3. COVID-19 new cases are predicted to increase 881 when the new cases from Addis Ababa city increase by 10,000 tests while holding other variables constant.

In addition, it is predicted to be 0 (negative cases not applicable) when the three variables are zero.

In the model, R^2^ = 96% of the variation in COVID-19 new cases was explained by the model (predictors). In the hypothesis test (F-statistic = 205.1 with DF = 3 & 26, p-value = 0.000), there was enough evidence to reject the null hypothesis that all the model’s coefficients are 0. The residual standard error = 12.16 shows how far the observed total COVID-19 new cases (Y-values) are from the predicted total COVID-19 new cases (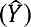)(Figure 7, R-software output).

**Figure 7:**
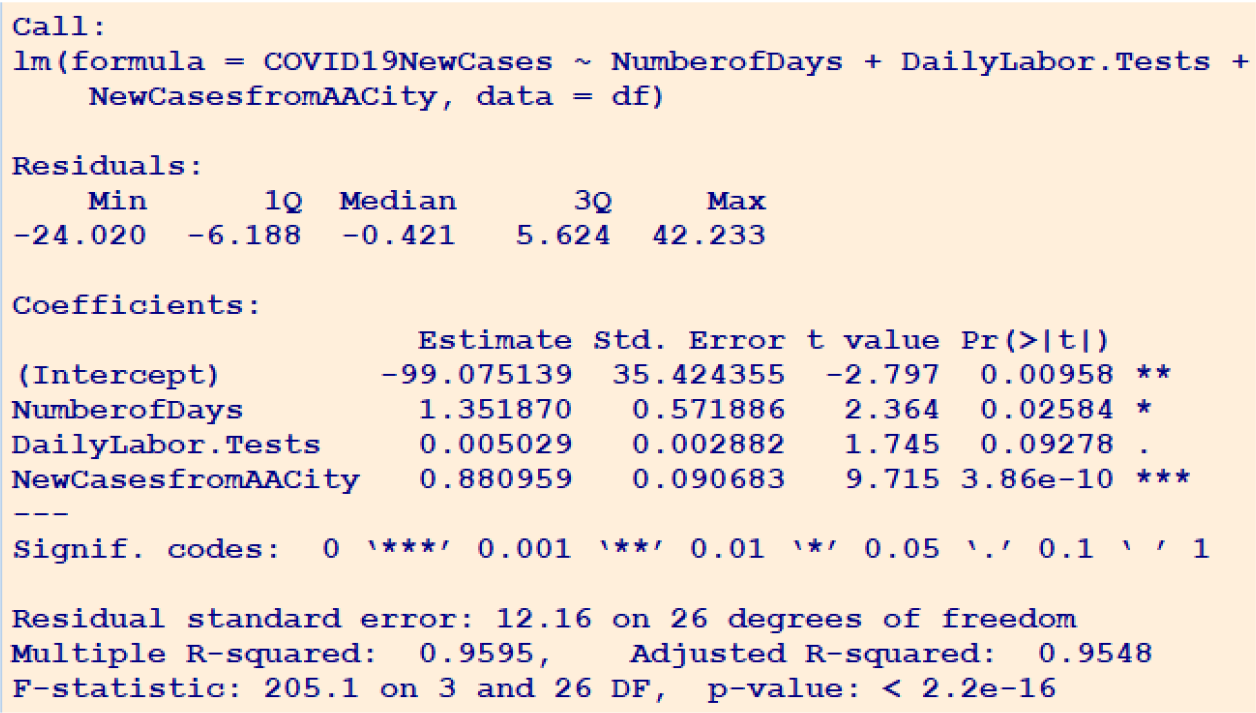
R Output of MLR for estimating the parameter to predict COVID-19 New Cases

### 3.9. Multiple Linear Regression (MLR) Model for New Deaths Due to COVID-19

In this model, new deaths due to COVID-19 were predicted significantly by the number of days and new recoveries at the 10% and 1% levels of significance, respectively.

The fitted MLR for new deaths due to COVID-19,

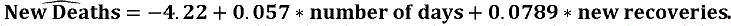

1. New deaths due to COVID-19 are predicted to increase by 6 when the number of days increases by 100 days while holding other variables constant.
2. New deaths are predicted to increase 79 when the new recoveries rise by 1,000 tests while holding other variables constant.
3. In addition, it is predicted to be 0 (negative deaths not applicable) when the two variables are zero.

In the model, R^2^ = 62.4% of the variation in new deaths was explained by the model (predictors). In the hypothesis test (F-statistic = 22.4 with DF = 2 & 27, p-value = 0.000), there was enough evidence to reject the null hypothesis that all the model’s coefficients are 0. The residual standard error = 1.07 shows how far the observed new deaths (Y-values) are from the predicted new deaths 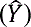(Figure 8, R-software output).

**Figure 8:**
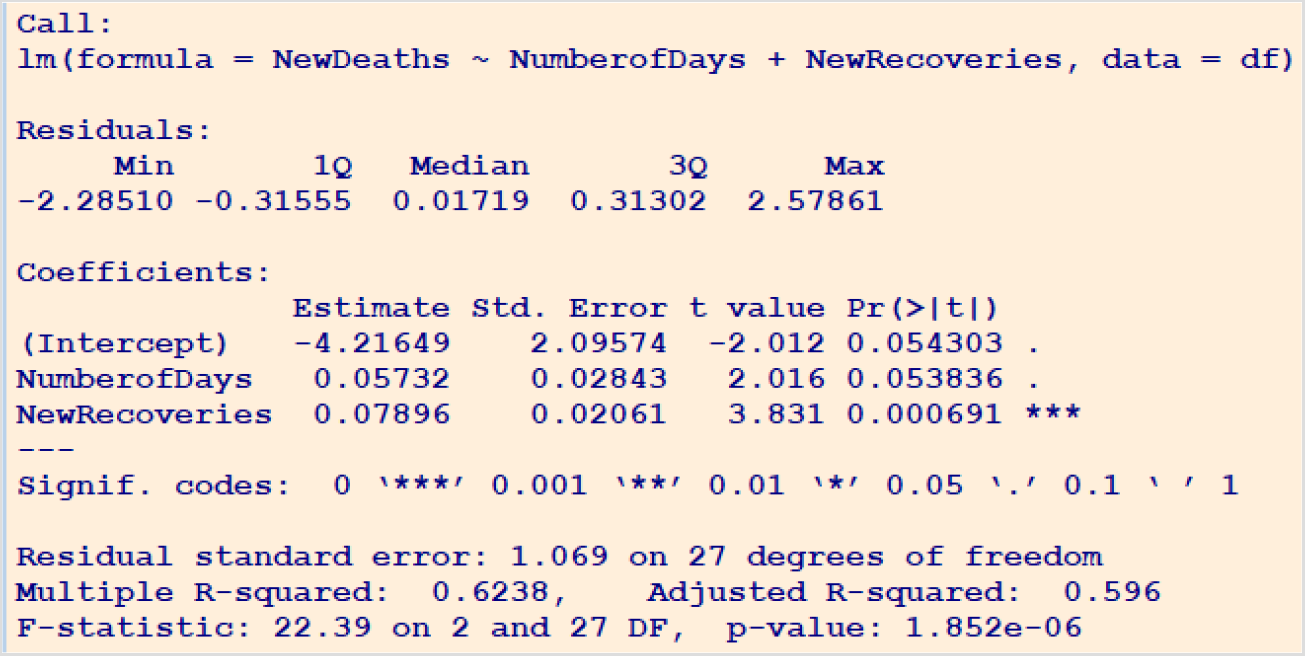
R-Output of MLR for estimating the parameter to predict new deaths

**Figure 9:**
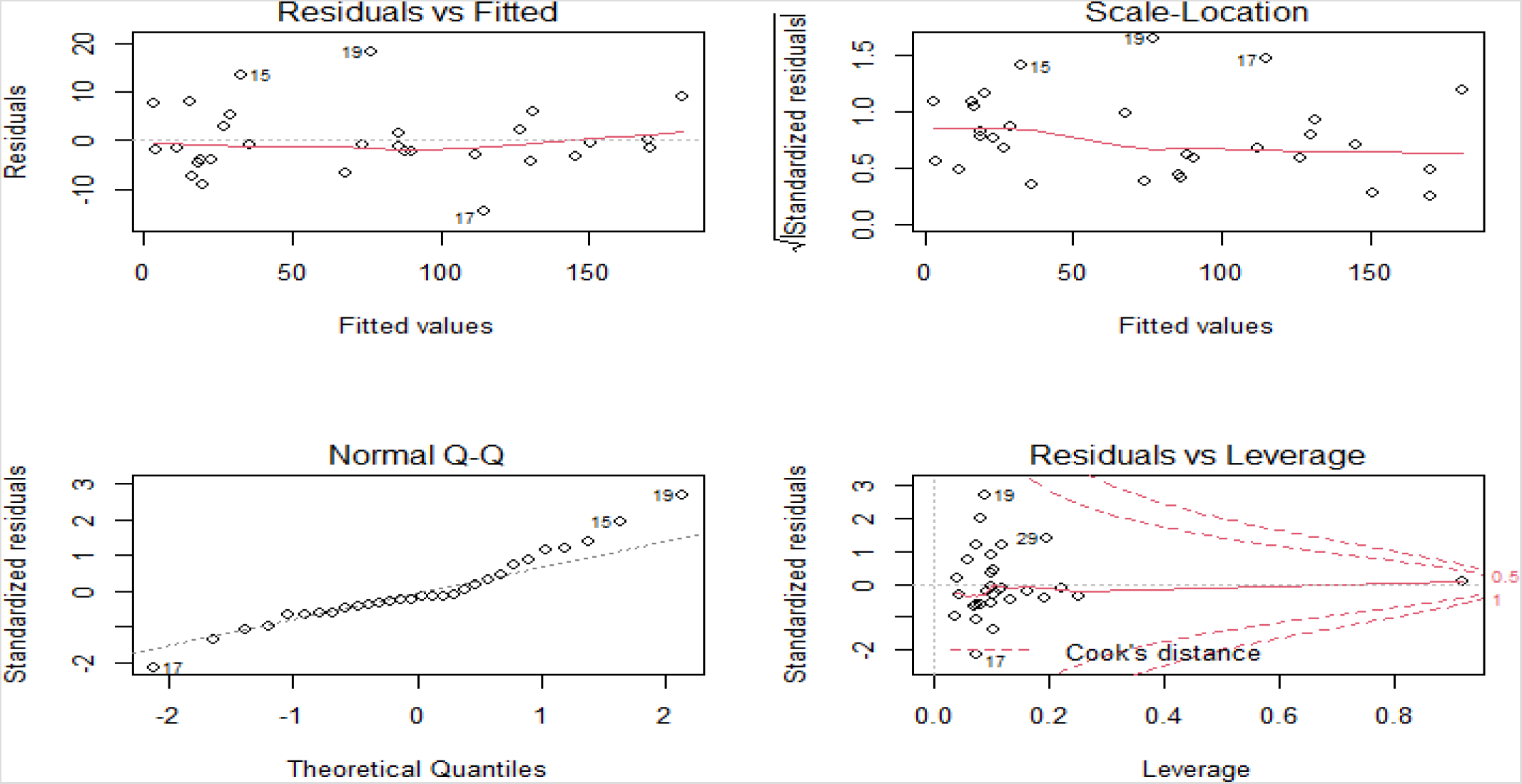
R-Output of Multiple Linear Regression Assumptions

## 4. Discussion

In the correlation analysis for COVID-19, new cases had significant and positive correlations with the number of days (r = 0.901), daily laboratory tests (r = 0.641), new recoveries (r = 0.389), new cases from males (r = 0.985), new cases from females (r = 0.964), new cases from AA city (r = 0.965), new cases from foreign natives (r = 0.416), new cases had unknown contact and travel histories (r = 0.958), and new cases had contacts with infected persons (r = 0.534).

In the correlation analysis, new deaths due to COVID-19 were significantly and positively correlated with the number of days (r = 0.648), COVID-19 new cases (r = 0.555), daily laboratory tests (r = 0.445), new cases from AA city (r = 0.533), new recoveries (r = 0.753), new cases of male (r = 0.53), new cases of female (r = 0.562) and the maximum age of new cases (r = 0.400, weak). However, it was significantly and negatively correlated with the minimum age of new cases (r = –0.426). As worldwide, a one study shown that there was a strong significant relationship between new deaths and new cases (r = 0.766, p = 0.01) per country. And, a similar study from Philippine showed that there was a correlation significantly between new cases and deaths (r = 0.440, p = 0.002). Other study from Algeria found there dates had correlations with daily new COVID-19 cases, and with daily new deaths [10–12].

And other highly qualified study from Indian found that the number of daily deaths was significantly and positively correlated with the number of recoveries (r = 0.28, p = 0.042), and the number of tests was significant and positively with COVID-19 cumulative infected cases (r = 0.59, p = 0.000) and with cumulative deaths (r = 0.55, p = 0.000). And, daily infections had significantly and positively correlated with daily deaths (r = 0.49, p = 0.000) [13].

The simple linear regression model was a better fit the data of COVID-19 new cases and new deaths than quadratic and cubic regression models. In this fitted model, COVID-19 new cases were significantly predicted by the number of days (*B* = 5.85), daily laboratory tests (*B* = 0.034), and new cases from Addis Ababa city *(B* = 1.2) at 5% level of significance. And, new deaths were significantly predicted by the number of days (*B = 0.12*), new cases *(B* = 0.016), and new recovered (*B = 0.1*) at 5% level of significance. A study from Indian found that the linear regression growth model was more specific to predict the number of affected cases of COVID-19 than the exponential growth model. These models used for forecasting in long term intervals. And, other study from Indian showed that the linear model was the best fitting model for Region III from May 3^rd^ to May 15^th^. Another study from India used simple linear regression analysis of the number of deaths as a function of the number of confirmed cases. In this study, the coefficient of determination (R^2^) was calculated to be 0.997, which implies a strong linear correlation between confirmed and dead cases. Whereas, the quadratic model was the best fitting model for Region II from April 6^th^ to May 2^nd^ [14–16].

In the multiple linear regression modl, COVID-19 new cases were predicted significantly by the number of days, daily laboratory tests and new cases from Addis Ababa city at the 5%, 10% and 1% levels of significance, respectively. Thus, COVID-19 new cases are predicted to increase 135, 503 and 881 when the number of days increases by 100 days, the daily laboratory tests increase by 100,000 tests, and the new cases from Addis Ababa city increase by 10,000 tests while holding other variables constant.

In the multiple linear regression model, new deaths due to COVID-19 were predicted significantly by the number of days and new recoveries at the 10% and 1% levels of significance, respectively. Thus, new deaths due to COVID-19 were predicted to increase 6 and 79 when the number of days increased by 100 days and the new recoveries rose by 1,000 tests while holding other variables constant, respectively.

This study agreed with a study from Kenya showing that there was a correlation between COVID-19 new cases and contact persons made by the confirmed status as well as the number of flights from foreign countries to Kenya. The study using univariate analysis of the generalized linear model showed that contact persons in Kenya had 0.265 effects on COVID-19 cases in Kenya. In the multivariate analysis, the contact persons and flights to Kenya had 0.278 and 3,309 effects on COVID-19 cases in Kenya at the 5% and 10% levels of significance, respectively. Researchers in Kenya also used the compound Poisson regression model, which showed that as the COVID-19 day increased by 235, the COVID-19 new cases were projected to 83,418 new cases [17].

A study from India used a linear regression analysis to predict the average week 5 and 6 death counts. In the study, there was a strong correlation between weeks 5 and 6 death counts with total cases, active cases, recoveries, and week 4 death counts. Despite this, the week 4 variables (total cases, active cases, and recoveries) were not significantly predicted by weeks 5 and 6 deaths count. However, the week 4 death counts significantly predicted the week 5 death count. Therefore, my study agreed with this study on the correlation analysis but not on the linear regression analysis [18].

### Limitations of The study

The main limitation of this analysis was that the data were not found together as collectively for all the previous reports and were taken from the face book and telegram pages of Ethiopia Ministry of Health. Second, limiting my analysis was that some data values were missed to report for 8 dates (such as the contact and travel history of the cases).

### Strength of the Study

Despite all the limitations, the greatest strength of this study was the very high adjusted R^2^ found in the predictive model. Three predictors for COVID-19 new cases were found in the multiple linear regression model, and its assumptions were fitted. In addition, there was cross-validation with two different software programs (R and SPSS).

## 5. Conclusion and Recommendation

### 5.1. Conclusion

There were 2,506 total COVID-19 cases and 35 deaths due to COVID-19 with a crude mortality of 1.4% from 14th march to 10^th^ June in Ethiopia, 2020. However, the total cases and total deaths of COVID-19 were 10 times and 6 times more, respectively, from the 12^th^ of May to the 10^th^ of June compared to the 14th of March to the 11^th^ of May 2020 in Ethiopia.

In the correlation analysis, the COVID-19 new cases were significantly correlated with the number of days, daily laboratory tests, new recoveries, new cases of males, new cases of females, new cases from Addis Ababa city, new cases with unknown contact and travel histories, and new cases that had contacts with infected persons.

In the correlation analysis, the new deaths due to COVID-19 were significantly correlated with the number of days, daily laboratory tests, COVID-19 new cases, new recoveries, new cases of males, new cases of females, new cases from Addis Ababa city, maximum age of new cases, and minimum age of new cases. In the simple linear regression, variables such as the number of days (coefficient *B* = 5.9), daily laboratory tests (coefficient *B* = 0.0334), new recoveries (*B* = 1.83), new cases of males (*B* = 1.58), new cases of females (*B* = 2.4), new cases from Addis Ababa city *(B* = 1.2), new cases from foreign natives (*B* = 19.2), new cases that had contact with other infected persons *(B* = 2.38), and new cases with unknown contact and travel histories *(B* = 1.2) significantly predicted COVID-19 new cases.

In the simple linear regression model, variables such as the number of days (*B = 0.124*), COVID-19 new cases *(B* = 0.016), daily laboratory tests (*B = 0.001*), new recoveries (*B = 0.104*), new cases from Addis Ababa city (*B = 0.019*), new cases of males (*B = 0.025*), new cases of females (*B = 0.041*), minimum age of new cases (*B = –0.104*) and maximum age of new cases (*B = 0.037*) significantly predicted new deaths.

In the multiple linear regression model, variables of the number of days, daily laboratory tests and new cases from Addis Ababa city significantly predicted COVID-19 new cases at the 5%, 10% and 1% levels of significance, respectively. Thus, COVID-19 new cases are predicted to increase 135, 503 and 881 when the number of days increases by 100 days, the daily laboratory tests increase by 100,000 tests, and the new cases from Addis Ababa city increase by 10,000 tests while holding other variables constant.

In the multiple linear regression model, variables of the number of days and new recoveries are predicted new deaths due to COVID-19 at the 10% and 1% levels of significance, respectively. Thus, new deaths due to COVID-19 were predicted to increase 6 and 79 when the number of days increased by 100 days and the new recoveries rose by 1,000 tests while holding other variables constant, respectively.

Finally, according to this analysis, if strong preventions and action are not taken in the country, the predicted values of COVID-19 new cases and new deaths will be 590 and 12 after two months (after 9^th^ of August) from now, respectively.

### 5.2. Recommendation

Even if Ethiopia has taken strong measures, including complete lockdown of both its internal and external borders and announced the command posts and keeping social isolation for the last three months, the number of new cases and deaths due to COVID-19 new cases were highly increased day to day. The research has predicted the total number of COVID-19 new cases where it is easy to see how it is likely to progress in the future. The above information should help the government make plans on how to deal with pandemics, especially when dealing with the current situation in Ethiopia.

The prevalence of the disease and its crude mortality from 12^th^ May to 10^th^ June 2020 increased more and more, and Ethiopia might be one of the top countries from Africa by leading this Pandemic for the next months if very strong necessary measures will not be taken into consideration. The government must come up with more isolation beds, more trained health care professionals, and more mass education and campaigns with the aim of ensuring that the public has information about how to stop the spread of the virus. Let us consider that a huge population of the Ethiopia population lives in rural areas, more education as well as infrastructure must be done in the rural area to make preparations in case the COVID-19 finds its way more for the districts and the villages.

Moreover, the Ethiopia government, the Ministry of Health and Regional Governments (especially the AA city administrative and Somali region) should give more awareness and protections collaboratively for societies, and they should also open more COVID-19 laboratory testing health centers in different areas of the country to ensure that those health centers can test more persons as the number of days increases, and the number of new cases will be highly increased. With these preventive and curative measures, the severity of COVID-19 will be limited when compared to other countries, such as the USA, South Africa and Egypt, which are now leading in the number of new cases and deaths in the world and in Africa. This research work will be extended after looking for the spread of the disease instantaneously by using a comparison of linear regression and nonlinear regression models.

## Data Availability

The data are available if required.

## Abbreviations

AA: Addis Ababa city
CI: Confidence Interval
MLR: Multiple Linear Regression
SLR: Simple Linear Regression

## Data Availability

The data are available if required.

## Funding

The research work was not supported by anyone.

## Conflict of interest

The author declares that I have no competing interests.

## Author contributions

All the research’s sections were performed by A.S. Argawu. I was the only author of the research.

## Acknowledgments

The data was collected from the daily reports of Ethiopia Ministry of Health’s, and Ethiopian Public Health Institute’s face book pages and telegram addresses from 12^th^ May to 10^th^ June 2020. So, I acknowledged them too much.

(https://www.facebook.com/EthiopiaMoH).

## Notes

### Competing Interest Statement

The authors have declared no competing interest.

### Funding Statement

The research work was not supported by anyone. It was supported by the author only.

### Author Declarations

I provide that my research work doesn't need of IRB review.

